# Low-Field Combined Diffusion-Relaxation MRI for Mapping Placenta Structure and Function

**DOI:** 10.1101/2023.06.06.23290983

**Authors:** Paddy J. Slator, Jordina Aviles Verdera, Raphael Tomi-Tricot, Joseph V. Hajnal, Daniel C. Alexander, Jana Hutter

**Affiliations:** Centre for Medical Image Computing and Department of Computer Science, University College London, London, United Kingdom; Cardiff University Brain Research Imaging Centre, School of Psychology, Cardiff University, Cardiff, UK; School of Computer Science and Informatics, Cardiff University, Cardiff, UK; MR Research Collaborations, Siemens Healthcare Limited, Camberley, United Kingdom; Centre for the Developing Brain, School of Biomedical Engineering and Imaging Sciences, King’s College London, London, United Kingdom; Biomedical Engineering Department, School of Biomedical Engineering and Imaging Sciences, King’s College London, London, United Kingdom

## Abstract

**Purpose:** Demonstrating quantitative multi-parametric mapping in the placenta with combined T2*-diffusion MRI at low-field (0.55T).

**Methods:** We present 57 placental MRI scans performed on a commercially available 0.55T scanner. We acquired the images using a combined T2*-diffusion technique scan that simultaneously acquires multiple diffusion preparations and echo times. We processed the data to produce quantitative T2* and diffusivity maps using a combined T2*-ADC model. We compared the derived quantitative parameters across gestation in healthy controls and a cohort of clinical cases.

**Results:** Quantitative parameter maps closely resemble those from previous experiments at higher field strength, with similar trends in T2* and ADC against gestational age observed.

**Conclusion:** Combined T2*-diffusion placental MRI is reliably achievable at 0.55T. The advantages of lower field strength - such as cost, ease of deployment, increased accessibility and patient comfort due to the wider bore, and increased T2* for larger dynamic ranges - can support the widespread roll out of placental MRI as an adjunct to ultrasound during pregnancy.

## Introduction

The placenta is vital to the health of mother and baby during pregnancy, performing the role of all organs and enabling exchange of oxygen, nutrients, and removal of waste products [1,2]. Placental dysfunction is a primary cause of many common pregnancy complications, such as stillbirth, fetal growth restriction (FGR), and pre-eclampsia [3]. However, human placental development remains poorly understood due to the difficulty of direct in-vivo observation [4]. Non-invasive biomarkers of human placental structure and function during pregnancy are therefore of great interest and can ultimately contribute to new techniques suitable for the prediction, diagnosis, and monitoring of pregnancy complications. This is particularly timely as there are potential treatments emerging for complications such as pre-eclampsia and FGR [5], which will be most effective if diagnosis is early and specific.

Placental MRI is a promising technique for diagnosis, prognosis and monitoring of multiple pregnancy complications including fetal growth restriction (FGR) [6] and pre-eclampsia (PE) [7]. In particular, T2* relaxometry is a promising technique for detecting pregnancy complications, with T2* reduced in FGR and PE [8,9]. The apparent diffusion coefficient (ADC) derived from diffusion MRI (dMRI) also shows promise as a biomarker, with lower ADC values in FGR [10–12]. The more complex dMRI Intravoxel incoherent motion (IVIM) model is also sensitive to pregnancy complications [13–17]. Recent *combined diffusion-relaxation* approaches merge relaxometry and dMRI into a single scan that accounts for correlations and between relaxation and diffusion properties [18] and can hence disentangle multiple complex placental microenvironments. Combined diffusion-relaxation has been demonstrated for T2-diffusion [19] and T2*-diffusion [20,21] in the placenta, and has shown promise for detecting a range of pregnancy complications at 3T [9,22,23].

Although placental MRI shows promise as a screening tool, there are several drawbacks that hinder widespread adoption. These include the high cost and time requirements associated with MRI scanning compared to ultrasound. This is especially pertinent for quantitative techniques such as T2* and dMRI, which require the acquisition of multiple volumes. Additionally, combined diffusion-relaxation MRI necessitates longer protocols than single T2* and dMRI scans, further exacerbating the issue. Furthermore, these techniques are typically based on single-shot Echo Planar Imaging (EPI) which, due to its low bandwidth, is sensitive to field inhomogeneities, such as at air-tissue boundaries.

Low-field MRI offers multiple advantages over high-field [24], and hence has the potential to enable widespread use of placental MRI. In particular, low-field MRI is generally cheaper to operate and easier to deploy; it can hence widen access to antenatal MRI beyond specialist centres. It allows a wider bore - expanding accessibility to the growing pregnant population with obesity - whilst maintaining field homogeneity, hence offering less susceptibility-induced distortions. The increased space within a wide bore, alongside lower acoustic noise, can also increase patient comfort. Furthermore, the longer T2* relaxation time, which results in a larger dynamic range, is a significant advantage, especially considering the low T2* values that are typically observed in late gestation and pathological cases.

In this paper, we show the feasibility of low-field quantitative placental MRI, using combined T2*-diffusion as an example. We first describe how we adapt a previously demonstrated multi-echo gradient-echo diffusion-weighted single-shot EPI sequence to a commercially available low-field scanner. We then calculate quantitative parameter maps from the low-field data and show qualitative agreement with those derived at higher field strengths. Our first demonstration motivates broader studies of low-field quantitative placental MRI for widespread pregnancy monitoring.

## Methods

We performed 68 scans on pregnant participants on a 0.55T clinical low field scanner (MAGNETOM Free.Max, Siemens Healthcare, Erlangen, Germany) after informed consent was obtained (MEERKAT project, REC 19/LO/0852). Participants were scanned in supine position maintaining frequent verbal interaction and life monitoring including heart rate and blood pressure readings. Before scanning each participant with our new low-field sequence, we performed an initial localizer scan, T2-weighted anatomical imaging, T1 relaxometry and short clinical diffusion sequences. Out of the 68 scans, 11 were discarded from further analysis due to an immediately visible contraction (9 scans) or a clear gross visual artefact (2 scans), leaving 57 scans.

This 57 scan cohort comprised 50 distinct participants, with seven participants who were scanned twice over gestation. The mean gestational age (GA) at scan time for the final 57 scan cohort was 29.9*±*5.84 [18.4,41.3] weeks, maternal BMI 28.1*±*4.84 [18.6, 37.6] kg/m2 and mean maternal age 34.3*±*5.19, [24.1, 44.5] years. In this initial study, we aimed to gain an initial insight into quantitative MRI at low-field by recruiting participants with a wide range of pregnancy complications and arranging them into broad cohorts. Specifically, participants were split into a healthy control cohort (30 participants who underwent a total of 36 scans) and three cohorts with pregnancy complications. These non-healthy cohorts are:

1. *Placental complication*: participants with significant multifaceted complications involving the placenta including molar pregnancies, prolonged premature rupture of the membranes and pre-eclampsia (10 participants who underwent 10 scans),
2. *Brain pathology:* fetal neurological pathologies, with no additional complications (9 participants who underwent 9 scans)
3. *T1 diabetes:* participants with type 1 diabetes but no additional complications (1 participant who underwent 2 scans).

We implemented a combined diffusion-relaxation multi-echo T2*-diffusion scan, using an adapted version of the ZEBRA sequence [25] as previously demonstrated at 3T [23]. We only used automatic shimming, with no specialist tools such as image-based shimming, which are required at 3T [26]. We scanned with two diffusion-weighting schemes, which we denote protocol 1 and protocol 2. For protocol 1, the b-values are 0, 50, 100, 150, 300, 500, 750, 1000 s/mm^2^. For protocol 2, the b-values are 0, 20, 30, 50, 70, 100, 120, 150, 300, 500, 750, 1000 s/mm^2^. For both protocols, each b-value was acquired with three echo times (TEs), 117, 161, 205 ms, and three orthogonal gradient directions were acquired for each non-zero b-value-TE pair. Compared to our previous 3T protocols [27], the highest b-value is lower and the TEs longer. These adaptations were made to account for the lower SNR and the reduced gradient performance available. The number of b-values and TEs were intentionally kept low to ensure the examination is widely available and accessible. Furthermore, due to the available implementation (i.e. not linked to the field strength), the lowest b-value (20 s/mm^2^) is 4 times higher than previously achieved at 3T [23], potentially reducing sensitivity to very high diffusivities associated with perfusion. No B0 map and calibration scans were acquired and no image-based shimming was performed, reducing both the scan time and need for expert knowledge. Scans were acquired in coronal orientation with respect to the mother with a 6-channel surface coil and in-built 9-channel table coil. Other acquisition parameters were: FOV = 400×400×1600mm, 4×4×4mm resolution, Grappa 2, TR = 11.1 s, partial Fourier 7/8. Protocol 1 has 66 total volumes and acquisition time 7 minutes 20 seconds. Protocol 2 has 102 total volumes and the acquisition time is 11 minutes.

The data was denoised using the mrtrix [28] implementation of MP-PCA [29] and direction-averaged for each b-TE combination. We analysed the direction-averaged data using a modified version of the dmipy toolbox [30] with a joint T2*-D model given by

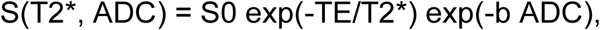

where TE and b are the acquired echo times and b-values respectively, T2* and ADC the obtained T2* and diffusivity values, and S0 is the signal at the b=0 volume with lowest TE. For the protocol 1 data we fit the model using the full dataset, i.e. all b-TE combinations. For the protocol 2 data, we fit the model to the full dataset, and also to the subset of the protocol corresponding to the protocol 1 b-TE combinations. We therefore obtained 57 protocol 1 T2* and ADC maps (i.e. all scans), and 28 protocol 2 T2* and ADC maps.

We also acquired a separate standard multi-echo gradient echo T2* scan in all participants, as previously reported [31]. The resolution was 3 mm^3^ isotropic, FOV = 400×400×[30-55] mm^2^ (slices were added or removed as necessary to maximise placental coverage), TR = 9670 ms and the TEs were 80 ms, 222 ms and 365 ms. The total acquisition time was 39 s. Of the 57 scans where we acquired combined T2*-diffusion data, 13 of the corresponding standard multi-echo gradient echo scans contained a clear contraction or visual artefacts and were hence removed, leaving 44 matched standard scans. We calculated T2* maps from these scans with a mono-exponential fit.

We manually defined a placenta mask and hence calculated the mean placental T2* and ADC from the joint T2*-ADC fit to the combined scan, and the mean placental T2* for the separate scans.

## Results

Figure 1 shows exemplary direction averaged data for a single scan after denoising. There is clear attenuation with increasing TE - with higher TEs revealing the lobular structure of the placenta. There is also clear signal attenuation with higher b-value, with the signal in the uterine wall attenuating quicker than the signal in the placenta.

**Figure 1:**
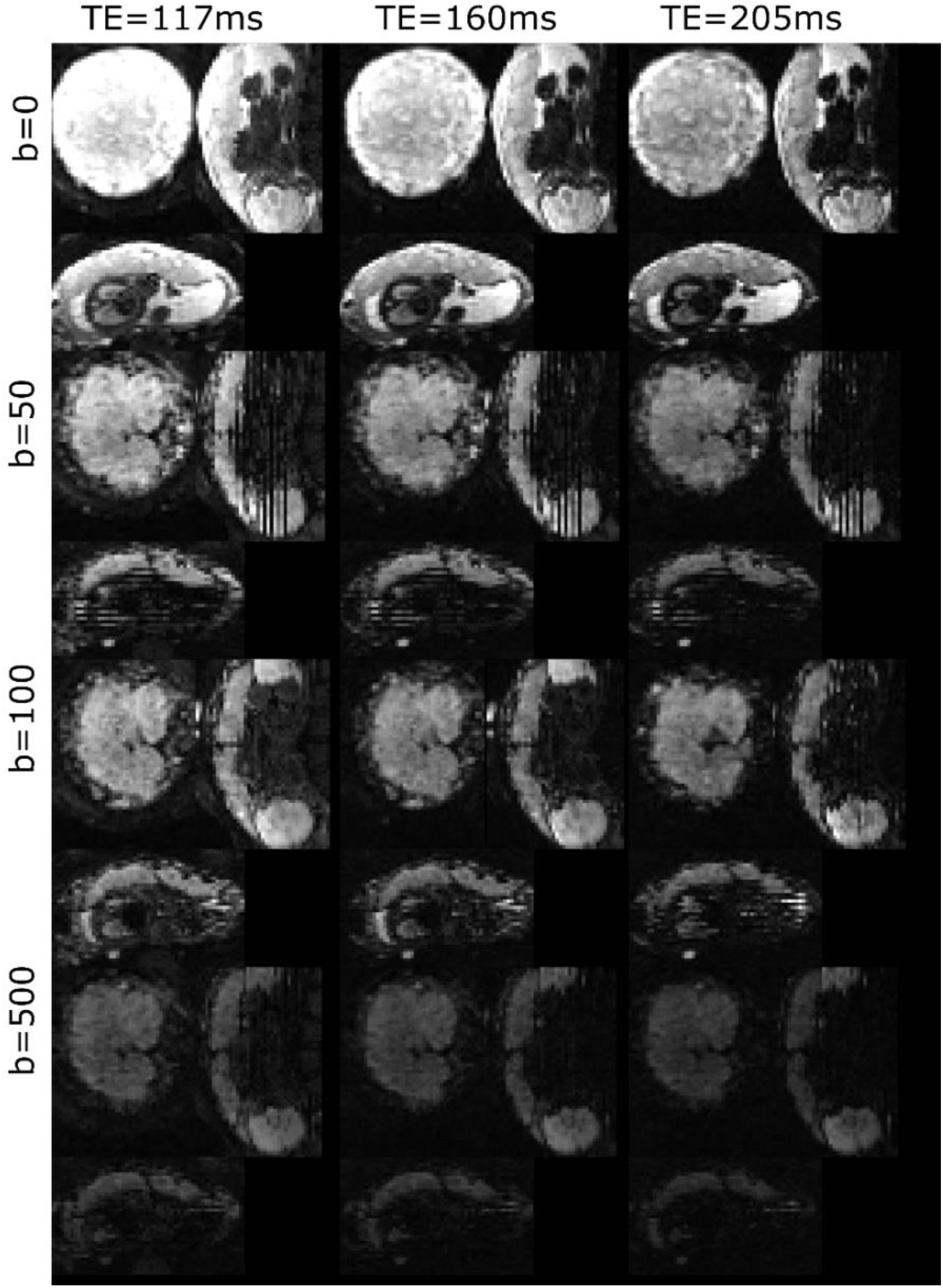
Exemplary direction-averaged data for three TEs and four b-values after denoising for a single control participant with GA = 27.14 weeks. See Figure 2 for derived maps for this participant.

Figure 2 shows example T2* and diffusivity maps from the T2*-ADC fit to protocol 1 data for the same participant as Figure 1. The maps clearly reveal placental features; there are lobular structures revealed in the T2* map, and higher diffusivity at the boundaries of the placenta.

**Figure 2:**
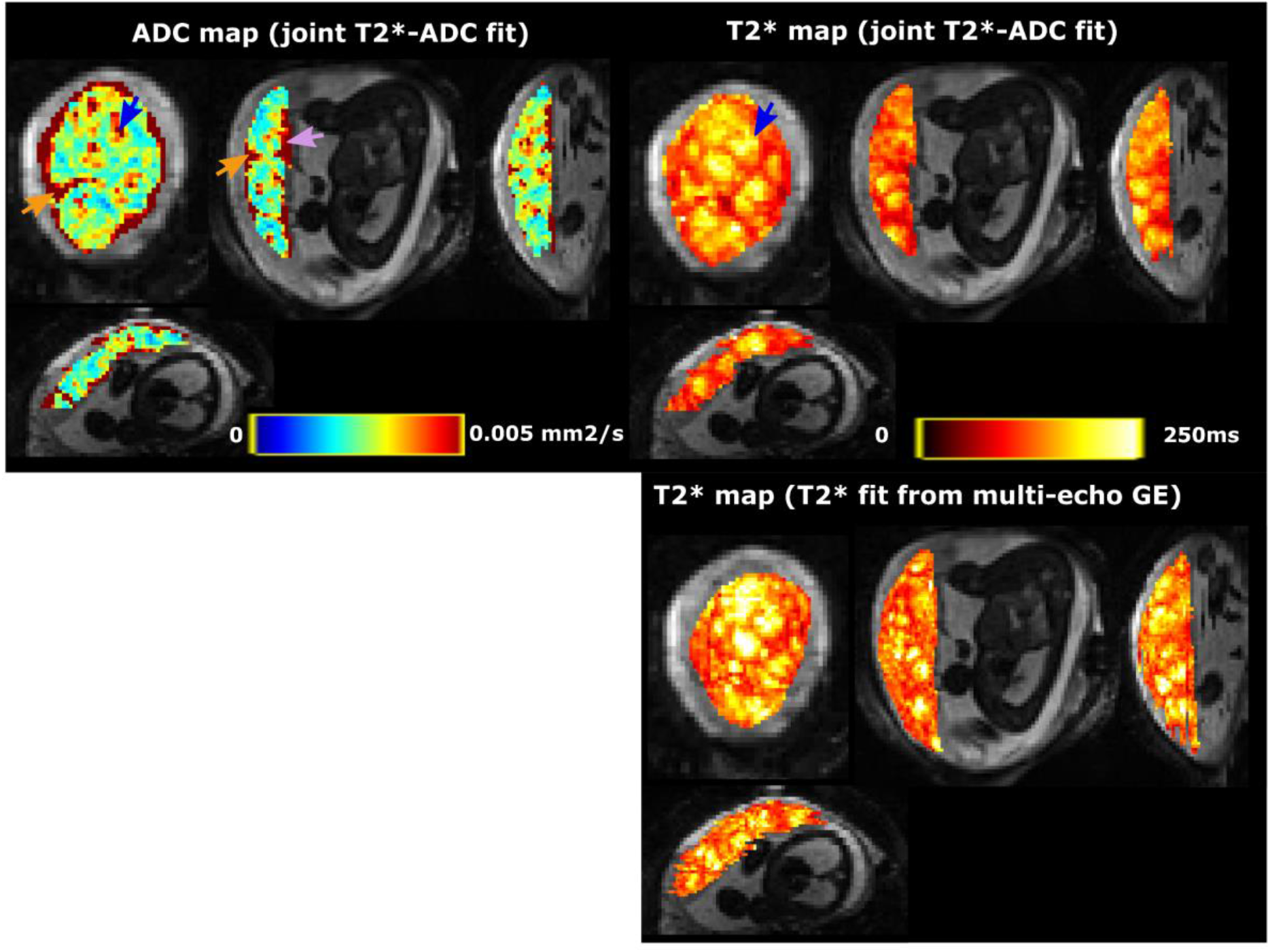
Detailed view into one example dataset (control, GA = 27.14 weeks - same participant as Figure 1). Top row: the ADC and T2* map from the joint fit. Bottom row: the T2* map from the additional multi-echo gradient echo scan. The blue arrow shows the increased ADC and T2* in the lobule centres, the pink arrow the increase on the chorionic plate and the orange arrows the increase in ADC and reduction in T2* in the septa between the lobules.

Figures 3 and 4 shows T2* and diffusivity maps for the T2*-ADC fits to the protocol 1 data, ordered by gestational age. There is a noticeable decrease in T2* values over gestation, whereas for ADC no clear pattern emerges. Figures 5 and 6 show the same maps for the subset of scans where protocol 2 data was acquired.

**Figure 3:**
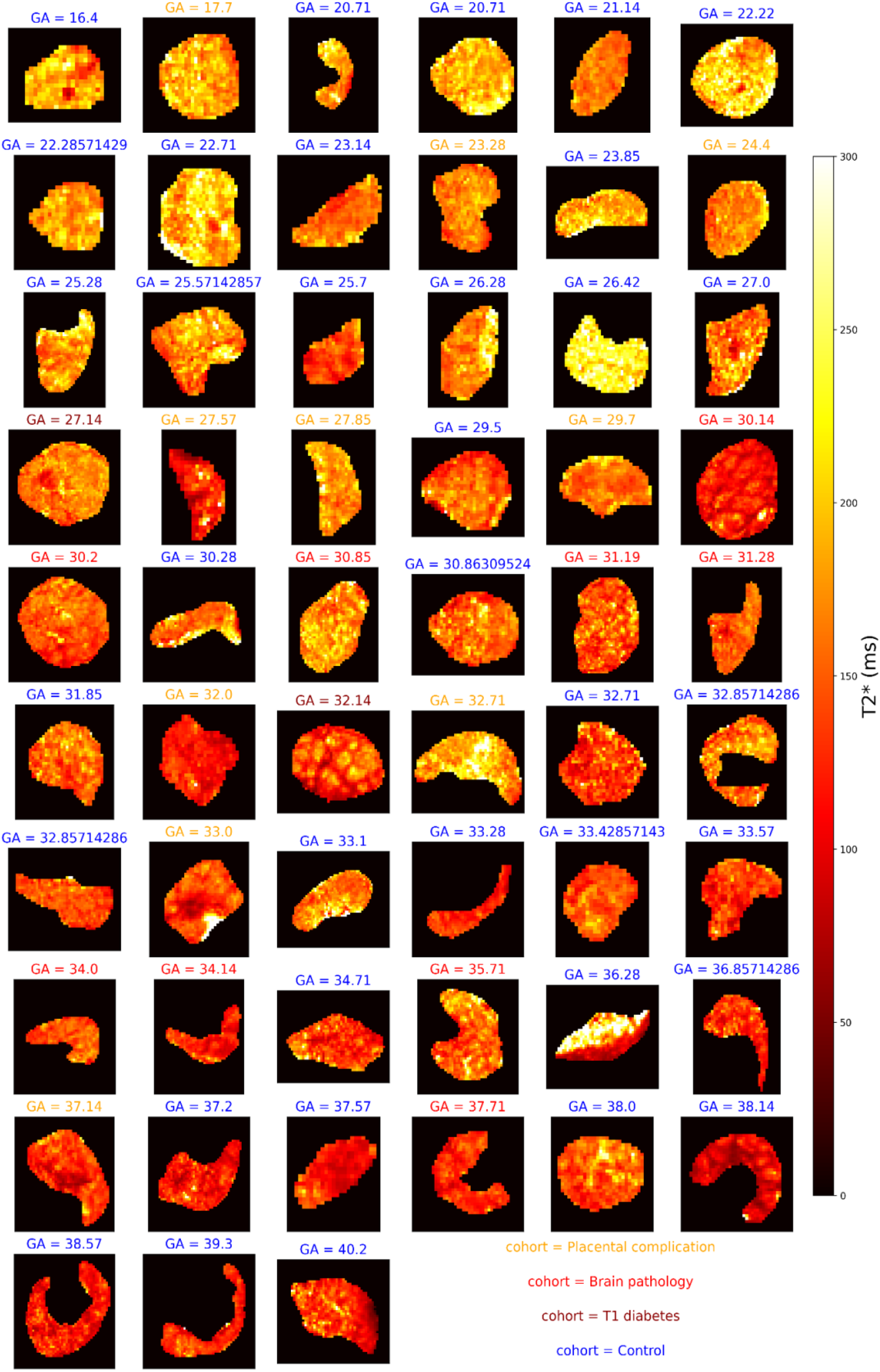
T2* maps across gestation from joint T2*-diffusivity model fit to the protocol 1 combined T2*-diffusion data. Subplot titles give the gestational age in weeks and text color denotes the cohort.

**Figure 4:**
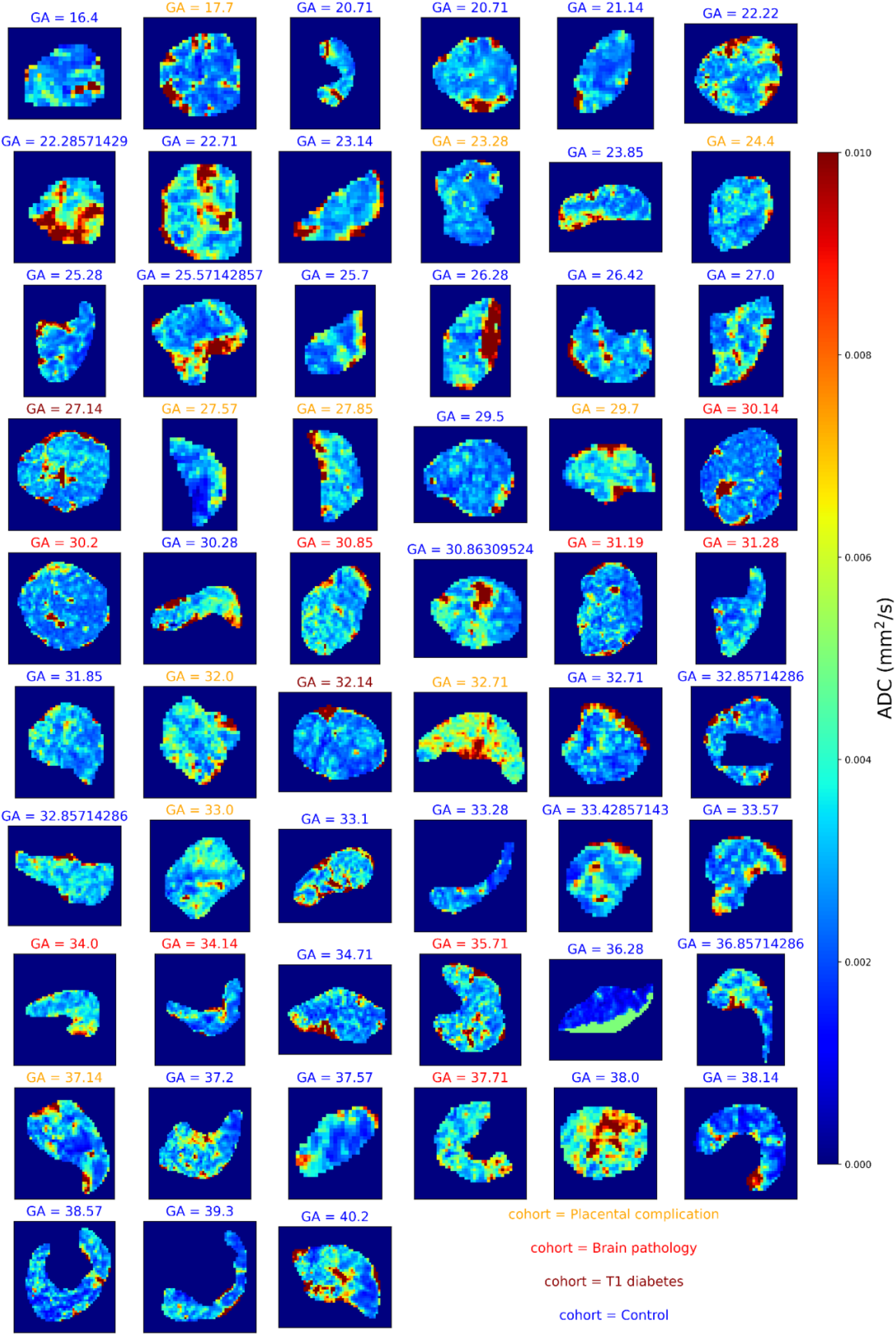
diffusivity maps across gestation from joint T2*-diffusivity model fit to the protocol 1 combined T2*-diffusion data.

**Figure 5:**
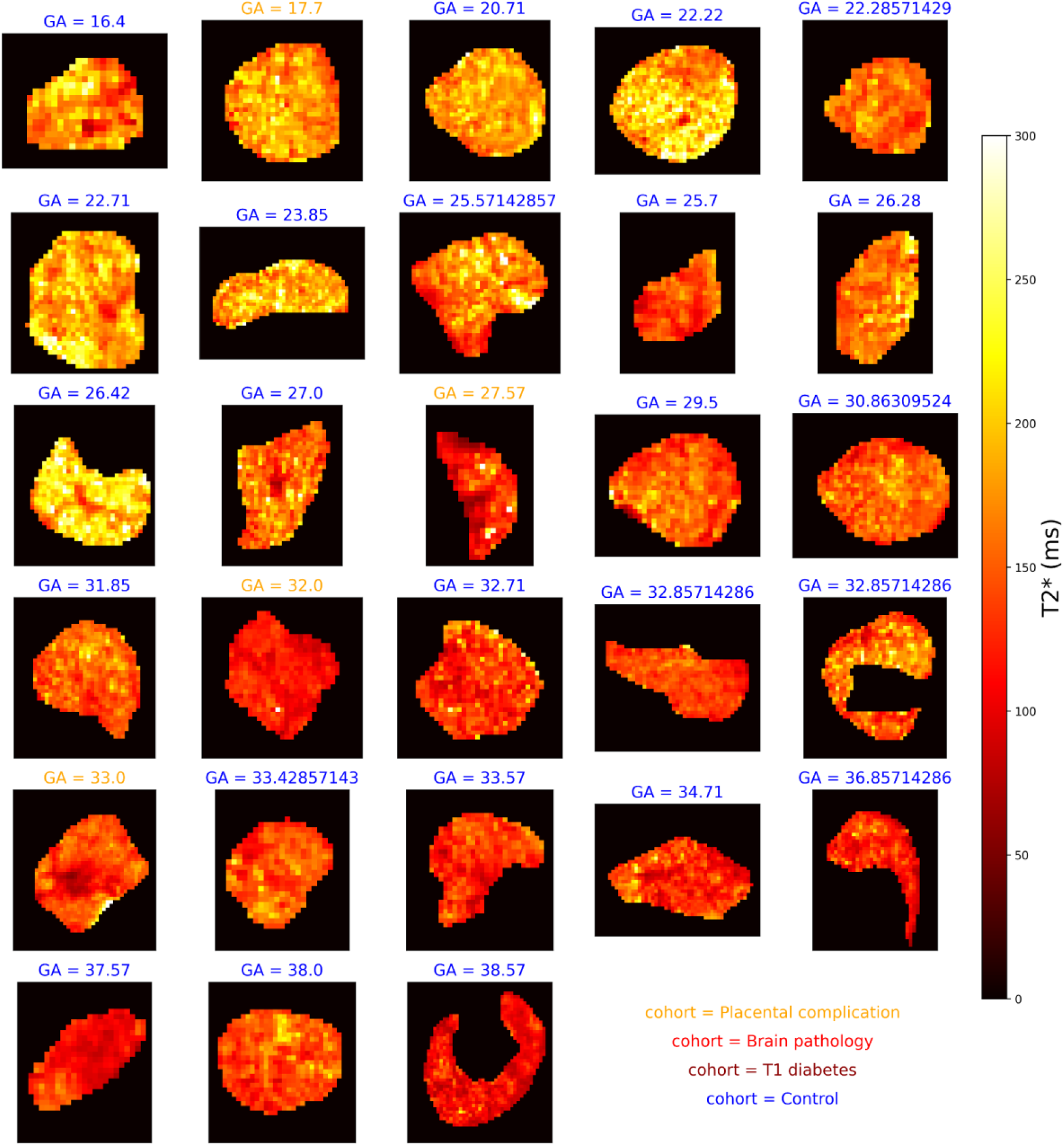
T2* maps across gestation from joint T2*-diffusivity model fit to the protocol 2 combined T2*-diffusion data.

**Figure 6:**
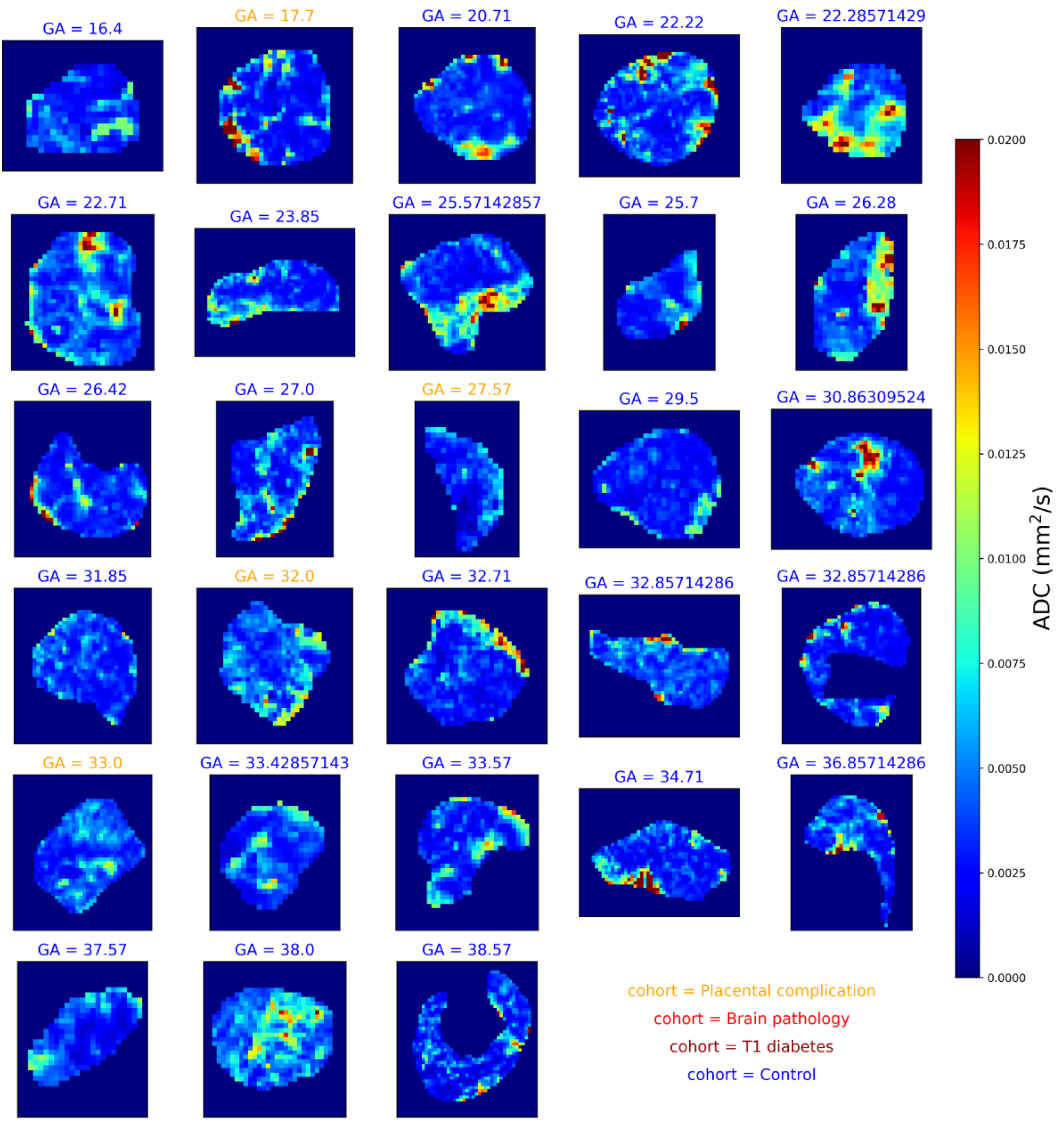
diffusivity maps across gestation from joint T2*-diffusivity model fit to the protocol 2 combined T2*-diffusion data.

Figure 7 shows the mean values of T2* (A) and ADC (B) in the placenta plotted against gestational age. There is a clear negative trend over gestation for T2* for both protocols, but no clear trend over gestation for the ADC. Figure 8 shows the differences in T2* and ADC when estimated from protocol 1 against protocol 2. The ADC estimated with protocol 2 is higher than with protocol 1, likely due to the additional low b-values. On the other hand, the T2* estimated with protocol 2 is lower than with protocol 1.

**Figure 7:**
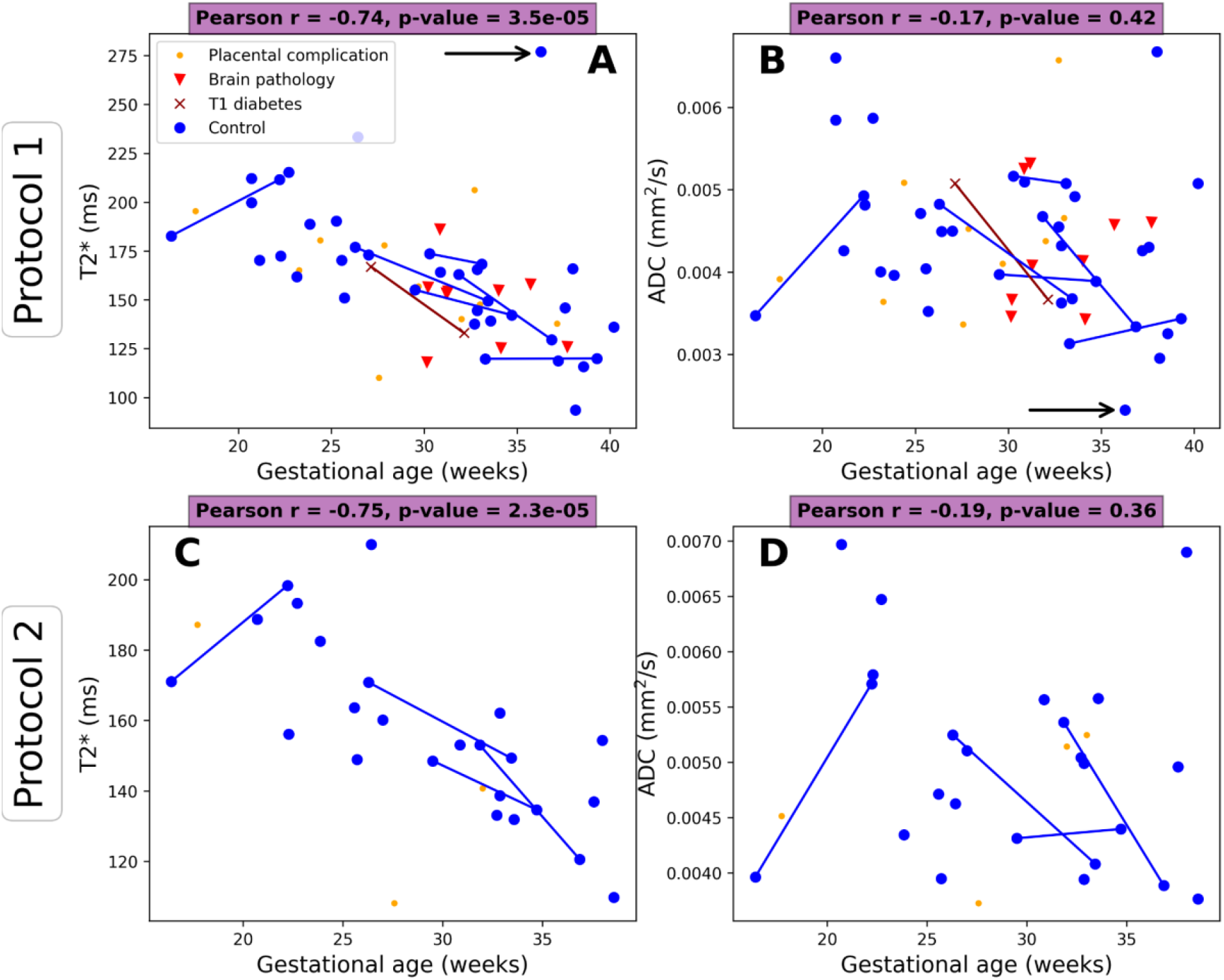
Mean T2* and ADC values over gestational age across the cohorts for protocol 1 (b=0, 50, 100, 150, 300, 500, 750, 1000 s/mm^2^) and protocol 2 (b=0, 20, 30, 50, 70, 100, 120 150, 300, 500, 750, 1000 s/mm^2^). Lines indicate the same participant scanned twice. Arrow in panel A indicates outlier scan with a clear contraction (see Discussion).

**Figure 8:**
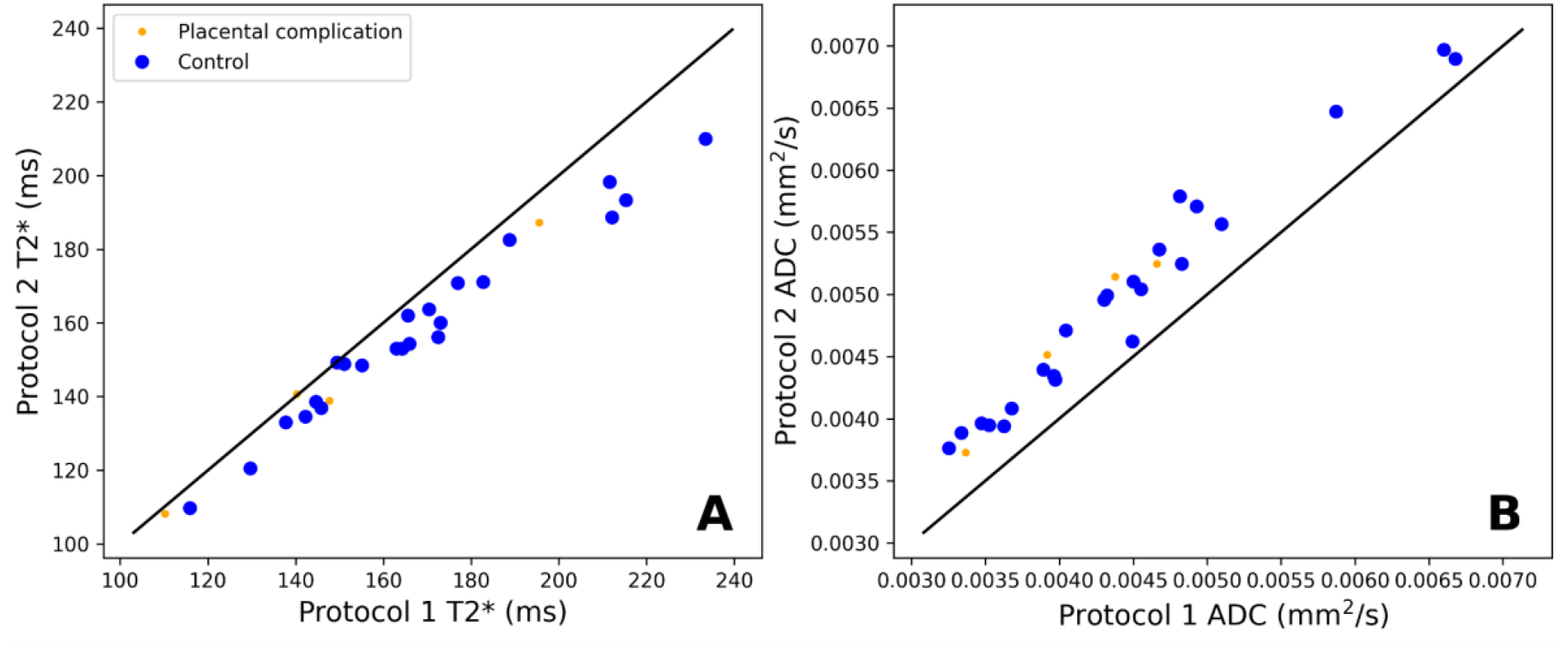
Estimated T2* (panel A) and ADC (panel B) values for both combined T2*-diffusion protocols.

Figure 9 shows mean T2* values against gestational age for the non-diffusion weighted multi-echo gradient echo scans and compares the mean T2* values between the combined T2*-diffusion and non-diffusion weighted scans. The left panel of Figure 9 reveals the same downward trend in T2* as observed in combined scans, with a slightly lower Pearson coefficient than the combined scan (−0.71 for the standard scan compared to -0.74 and -0.75 for the protocol 1 and protocol 2 combined scans respectively). The middle and right panels of Figure 9 shows that the standard scan routinely estimates higher T2* than the combined scans.

**Figure 9:**
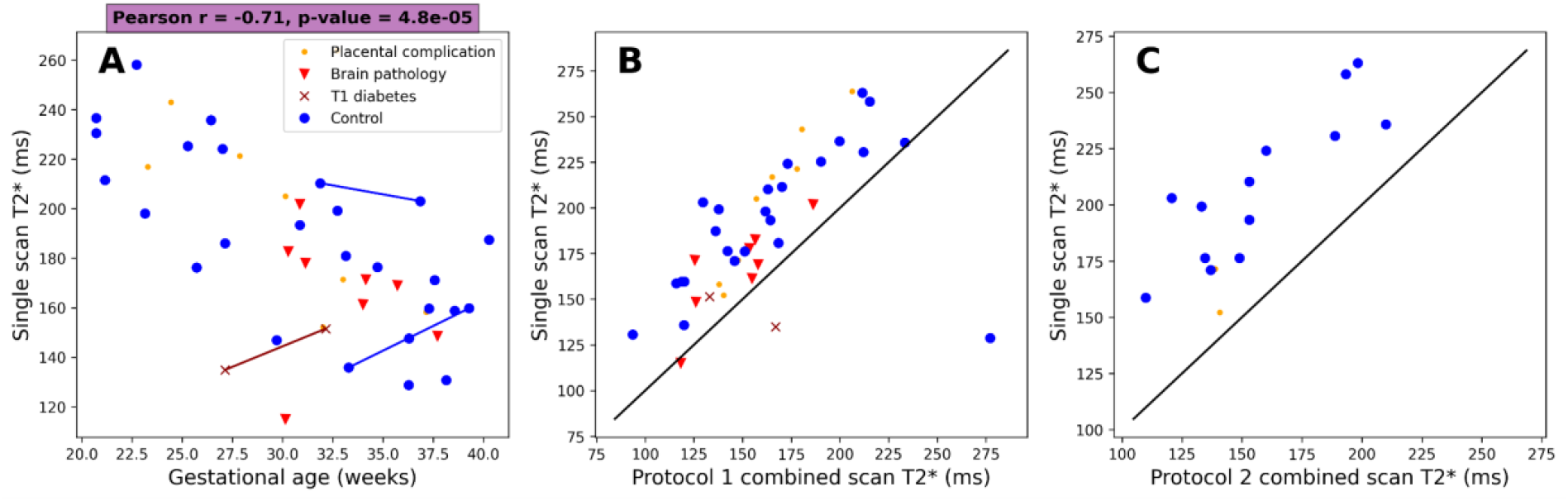
A) Mean T2* values against gestation from monoexponential T2* fit to standard multi-echo gradient echo scan. B) Mean placental T2* values from joint model fit to combined T2*-diffusion scan (protocol 1) against mean placental T2* values from fit to standard multi-echo gradient echo scan. C) As middle panel but for protocol 2.

## Discussion

Our study demonstrates quantitative placental MRI at 0.55 T, which offers several advantages over high-field scanning for the purpose of pregnancy monitoring. In addition to the potential for reduced cost and thus increased accessibility, low-field MRI can address specific issues related to pregnancy, such as a larger bore size allowing access to this modality for the growing number of overweight and women with obesity, and reduced distortion artefacts and B1 inhomogeneity, addressing some image quality challenges in abdominal/fetal MRI and foregoing the need for specialist shimming tools and expertise. Deriving quantitative metrics from low-field MRI has the potential to enable early and specific diagnosis of pregnancy complications, including pre-eclampsia and FGR, in a much broader range of settings than high-field MRI alone. This could significantly improve pregnancy monitoring and management, especially in low-resource settings.

On first inspection, our 0.55T scans contain similar information to previous work at 1.5T and 3T. Namely, the low-field T2* and diffusivity maps in Figures 2-6 have comparable patterns to those previously observed at higher field strengths, despite different scanning parameters such as the voxel size (4mm^3^ isotropic here compared to ∼2mm^3^ at 3T, e.g. [23]). Specifically, many T2* maps (Figures 2, 3, 5) reveal the lobular structure of the placenta, potentially revealing the oxygenation level of maternal blood, as previously observed in standard T2* [7,32,33] and combined T2*-diffusion [21,23] experiments. There is also a higher diffusivity at the boundaries of the placenta in ADC maps potentially reflecting areas with high volumes of maternal blood perfusing into the placenta (Figures 4 and 6). This was previously shown in diffusion [27] and combined T2*-diffusion [21,23,27] scans. These observations highlight that combined T2*-diffusion MRI at 0.55T is viable and promising.

As expected, the T2* values calculated from our low field scans are higher than those previously reported at higher field strengths. A directly comparable combined T2*-diffusion study at 3T showed T2* values of ∼80 ms at GA=20 weeks and ∼30 ms at term (GA = 40 weeks) [21]. This compares to our T2* values of ∼200 ms (GA=20 weeks) to ∼125 ms (at term) for both protocol 1 and protocol 2 (Figure 7).

For our standard, separate T2* acquisition, the T2* ranged from ∼250 ms at GA=20 weeks to ∼150 ms at GA=40 weeks (Figure 9, left panel). Again, as expected, this is much higher than directly comparable studies at higher field strengths. Specifically, to compare to the most recent studies of T2* at 3T, Schabel et al. showed T2* values ranging from ∼80 ms (GA= 20 weeks) to ∼30 ms (at term) [34], and Ho et al. had T2* ∼90 ms (GA = 20 weeks) to ∼30 ms (at term) [7]. Regarding 1.5 T, the recent study by Sinding et al. showed T2* ∼150 ms at 20 weeks to ∼60 ms at term [35]. The observed values are, however, consistent with a previous low field study at a dedicated EPI scanner [36].

Therefore, interestingly, despite having fewer echo times, we observe a comparable, or potentially wider, range in T2* values at 0.55T likely due to these longer relaxation times. This may allow for better quantification of more subtle and individual differences in oxygenation in pregnancy complications where T2* is reduced [8], by enabling wider separation of controls and cases.

We found that measuring T2* with a standard, separate T2* scan routinely estimates higher T2* than the combined scan (Figure 9, right panel). We also observed that the combined T2*-diffusion protocol with more b-values (protocol 2) resulted in higher ADC and lower T2* values (Figure 8). This likely reflects the different TEs in the combined (TE = 117, 161, 205 ms) and standard (TE = 80, 222, 365 ms) scans. Additionally, combined T2*-diffusion scans may inherently yield different values than separate T2* scans, due to a portion of the T2* decay being fit by the diffusion component of the model and vice-versa. The extent to which the separate and/or combined scans reflect the true, underlying, T2* value is an open question that warrants further investigation, by first undertaking combined and standard scans with matched TEs.

Despite removing datasets with obvious contractions before analysis, we examined the most prominent outlier (black arrows in Figure 4) and observed a clear contraction. This highlights the importance of viewing the data to look for outlier events such as contractions, as previously observed [37–39][37].

### Limitations and Future Work

The presented results hint that low-field maps display wider across-scan variance than high field. This could be because our preprocessing pipeline has been developed for less time. In future, we will continue building our pre-processing pipelines for low-field placental MRI by developing and integrating low-field specific methods for motion correction and artifact correction.

This study has reduced b-value coverage, particularly at very low b-values, to previous studies at higher field strength. For example, the minimum b-value in [27] was 5 s/mm^2^ compared to 20 or 50 s/mm^2^ here. This may lead to diffusivity maps with lower ranges than in comparable studies at 1.5 T and 3 T. An obvious path for further investigation is to collect repeat combined T2*-diffusion (and separate T2*) measurements for the same participants on the same day at 0.55 T, 1.5 T, and 3 T.

We assumed that T2* and diffusion decay are both monoexponential. This is suboptimal, particularly as it has been previously shown that multi-compartment models are required to adequately explain the diffusion MRI signal in the placenta [27]. In future we will explore such models, as previously demonstrated in T2-diffusion [19,22] and T2*-diffusion [21] experiments. Although in principle we could also explore multi-exponential decay models of the T2* signal, in practice this would likely require more TEs than the current three. As well as multi-compartment modelling, we will also explore data-driven unsupervised machine learning approaches that don’t require fixing the number of model compartments [40].

In this study, we scanned coronally to the maternal habitus, and displayed parameter maps of the largest slice through the placental parenchyma (in Figures 2-6). In future, specific visualisation and analysis of the processes from spiral arteries to the chorionic plate would be simplified by transforming the placenta into a common, biological space as previously proposed [41,42].

An obvious and compelling avenue for further research involves conducting a multi-centre study with a larger participant pool, including a wider range of fine-grained pregnancy-related complications, rather than the broad categorisation of complications we use here. This study would have increased statistical power and could identify more nuanced relationships between placental MRI metrics and specific pregnancy complications.

## Conclusion

We demonstrate placental quantitative MRI at 0.55T. Our results show many common features with studies conducted at higher field strengths. Our findings can support wider deployment of quantitative placental MRI as a complementary tool to ultrasound for pregnancy monitoring.

## Data Availability

All data produced in the present study are available upon reasonable request to the authors.

## Acknowledgements

We thank all mothers, midwives, obstetricians, and radiographers who played a key role in obtaining the datasets. Grant support: NIH (1U01HD087202-01); Wellcome Trust (201374/Z/16/Z); EPSRC (EP/V034537/1, EP/M020533/1); UKRI (MR/T018119/1 JH); MRC (MR/V002465/1); NIHR Biomedical Research Centre at UCLH NHS Foundation Trust and UCL; core funding from the Wellcome/EPSRC Centre for Medical Engineering at KCL (WT 203148/Z/16/Z); the NIHR Biomedical Research Centre based at Guy’s and St Thomas’ NHS Foundation Trust and KCL. The views expressed are those of the authors and not necessarily those of the NHS, the NIHR or the Department of Health.

